# Peripheral Biomarkers and Determinants of Survival for Patients with Melanoma Brain Metastasis Treated with Immune Checkpoint Blockade

**DOI:** 10.64898/2026.10.19.26365947

**Authors:** Jasmine Hassan, Martin Little, Andrea Farkas, Sophie MacKay, Nicholas Anthony Coupe, Mark Ross Middleton, Miranda Jayne Payne, Benjamin Peter Fairfax

**Affiliations:** Department of Oncology, University of Oxford, Oxford, United Kingdom; Weatherall Institute of Molecular Medicine, University of Oxford, Oxford, United Kingdom; Cancer and Haematology Centre, Oxford University Hospitals NHS Foundation Trust, Oxford, UK; NIHR Oxford Biomedical Research Centre, Oxford University Hospitals NHS Foundation Trust, Oxford, UK

**Author notes:** Contributing authors. These authors contributed equally and share first authorship (Jasmine Hassan, Martin Little).

**Keywords:** Melanoma, Brain Metastasis, Checkpoint Immunotherapy, Real World Data

## Abstract

**Background:** Immune checkpoint blockade (ICB) has become the standard of treatment for metastatic melanoma (MM). However, patients with melanoma brain metastases (MBM) are often excluded from clinical trials. Here we analyse patients within the real-world OxCITE cohort receiving ICB treatment for metastatic melanoma, with a focus on MBM, aiming to discern outcomes, prognostic factors and peripheral biomarkers.

**Methods:** Patients eligible for ICB treatment were recruited prospectively. Demographic, clinical and radiological data were collected from electronic patient records. Plasma samples were collected prior to the first cycle of treatment for plasma proteomic analysis.

**Results:** 307 patients were included, 81 of whom had MBM on commencing ICB. No difference in overall survival between patients with and without brain metastases was noted on multivariable analysis (HR=1.1, 95% CI 0.73-1.6, p=0.68), whereas hepatic metastases were associated with increased risk of death (HR 1.7, 95% CI 1.2-2.4, p=0.0019). Within the MBM group, steroid treatment (HR=2.5, 95% CI 1.3-5.0, p=0.006) was associated with poor OS, whilst treatment with single agent ICB (HR=1.9, 95% CI 0.86-3.9, p=0.11), symptomatic first presentation of MBM (HR=1.5, 95% CI 0.76-2.8, p=0.25) and anti-seizure drug treatment (HR=1.74, 95% CI 0.89-3.4, p=0.10) were not. Amongst all patients carriage of a *BRAF* mutation was associated with increased risk of MBM (OR 2.49, 95% CI 1.4-4.4, p=0.0007), and increased propensity to new brain metastases on ICB treatment (OR 2.5, 95% CI 1.1-5.9, p=0.036). Analysis of blood and plasma samples demonstrated that patients with MBM have altered haematological profiles, with raised NLR and increased plasma glial fibrillary acidic protein levels, reinforcing the distinct systemic features of MBM.

**Conclusions:** In this real-world cohort of patients receiving ICB for metastatic melanoma, the presence of MBM did not associate with worse OS. Conversely, baseline corticosteroid use was adversely prognostic. Intriguingly, plasma GFAP was significantly elevated in patients with MBM and correlated with the burden of intracranial disease, highlighting a potential role for GFAP as a candidate biomarker for MBM in high-risk surveillance. Our observations indicate the adversely prognostic weight historically attached to M1d disease is no longer applicable in the ICB era.

## 1 Introduction

Brain metastases generally portend poor survival across cancer types. In the United Kingdom, melanoma is the third most common cause of brain metastases[1], despite being the fifth most common cancer overall[2], with *>*80% of cases being early stage disease[3]. This skewing reflects the high central nervous system (CNS) predilection of melanoma. This is most evident in late-stage untreated disease, where 40-50% of stage IV melanoma patients progress to develop melanoma brain metastases (MBM)[4]. This exceptional tropism of melanoma to the brain has rendered presence of MBM an important treatment consideration in advanced disease. Furthermore, the historically negative association between CNS metastases and outcome led to the 8th edition of the American Joint Committee Cancer Staging Manual incorporating CNS metastases into a separate substage (M1d)[5]. Of note, melanomas carrying an activating *BRAF* mutation are reported to have a further heightened risk of MBM, although the consistency of this observation varies between centres[6, 7]. Although patients with active MBM have been excluded from clinical trials, including Checkmate 067[8], early clinical experience demonstrated response of MBM to immune checkpoint blockade (ICB)[9]. These observations have led to specific studies for patients with CNS disease, including Checkmate 204[10] and ABC[11], which have in turn shaped current guidelines. These trials underscored the robust activity of ICB towards MBM, with Checkmate 204 reporting 71.9% overall survival (OS) at 3 years in their asymptomatic cohort[10], and ABC reporting 3-year OS of 57% and 7-year OS of 48%[11]. However, trial data is not always reflective of real world experience, knowledge of which is vital to better inform patients of representative outcomes as well as being core to the identification of other determinants of response. Given this, we have analysed real-world outcomes of patients who were prospectively incorporated into the Oxford Cancer Immunotherapy Toxicity and Efficacy (OxCITE) cohort and who have received ICB therapy for metastatic melanoma in Oxford, specifically focusing upon outcomes of patients affected by MBM.

## 2 Methods

### 2.1 Participants

Patients eligible for ICB treatment of metastatic melanoma were prospectively recruited between 23rd November 2015 and 28th February 2025 at Oxford University Hospitals NHS Foundation Trust. Written consent was obtained from all patients to donate samples to the Oxford Radcliffe Biobank (Oxford Centre for Histopathology Research ethical approval reference 19/SC/0173, project nos. 16/A019, 18/A064 and 19/A114), and to grant access to their routine clinical data. No compensation was offered for this consent. Patients were treated with either combination ICB (ipilimumab 3 *mg kg^→^*^1^ plus nivolumab 1*mgkg^→^*^1^ every three weeks for *→* 4 cycles followed by maintenance nivolumab) or single agent ICB (pembrolizumab 200mg every three weeks or 400mg every six weeks, or nivolumab 240 mg every two weeks or 480mg every four weeks.

### 2.2 Clinical Outcomes

Demographic, clinical and radiological data were collated from electronic patient records. Presence of MBM was defined as at least one brain metastasis or leptomeningeal disease being reported on either MRI head with contrast or CT head with contrast preceding the first cycle of ICB treatment. Presence of hepatic metastasis was defined as at least one hepatic metastasis being reported on CT with contrast or MRI preceding the first cycle of ICB treatment. Haemorrhage associated with brain metastases was defined as symptomatic intracranial haemorrhage in a patient with MBM. Symptoms of brain metastases were defined as focal neurological symptoms, symptoms of raised intracranial pressure, acute confusion or seizures associated with a diagnosis of MBM. Steroids were defined as a prescription being issued for oral or intravenous corticosteroids which was active at the time the patient commenced ICB treatment. Anti-seizure medications were defined as those started for prophylaxis or treatment of seizures that continued to be taken at the start of ICB treatment. Local interventions for MBM prior to initiation of ICB were recorded for stereotactic radiosurgery (SRS), surgical resection, or a combination of both. Objective response was not available for this cohort. Overall survival (OS) was defined as time from treatment initiation to death from any cause.

### 2.3 Sample Collection

Baseline absolute lymphocyte count, absolute neutrophil count and lactate dehydrogenase (LDH) were obtained from electronic patient records, generated via Sysmex XN series analyzers. The blood sample taken closest to initiation of ICB, between day-30 and day 0, was used for analysis. Glial fibrillary acidic protein (GFAP) levels were measured via a nucleic acid-linked immuno-sandwich assay (NULISA, Alamar), using patient plasma diluted 1:2 in HBSS taken immediately pre-treatment on day 0[12]. GFAP measurement was represented as normalised protein quantification units (NPQ), a relative log2 scale where each unit increase indicates a doubling in concentration.

### 2.4 Statistical Analysis

Univariable analysis of continuous vs discrete variables was performed using Mann-Whitney U tests. Univariable linear regression was performed using Spearman rank correlation coefficient. Associations between discrete variables were analysed through univariable and multivariable binomial logistic regression models which, at baseline, included age, sex, *BRAF* status, performance status and pre-treatment with BRAF/MEK inhibitors (BRAF/MEKi) or ICB unless specified otherwise. *BRAF* status was included due to prior associations between *BRAF*-mutated disease and brain metastases[13, 14]. Exposure to prior BRAF/MEKi was similarly included due to previous associations with reduced response rate to combination ICB[15], whilst post-ICB exposure progression reflects immunotherapy-resistant disease, impacting OS and propensity to developing new brain metastases. Survival analysis was performed using univariable Cox regression which included the same variables, multivariable survival analysis incorporating covariates with p *→*0.1 in univariable analysis unless specified otherwise. Median follow up was calculated using the reverse Kaplan Meier approach. An *ω* of *→*0.05 was deemed significant, and we did not correct for multiple tests.

## 3 Results

### 3.1 Demographics of brain metastases

The study cohort included 307 patients with MM, of whom 81 patients had MBM upon inclusion within the cohort when commencing ICB treatment. The median follow-up for the whole cohort was 60 months (95% CI 48.6-68.3) with a total of 142 events, with a median follow-up of 36 months (95% CI 19.7-68.3) and 36 events for those with brain metastases. We found patients with MBM at the start of ICB were on average younger (median 62 years (IQR 53-72) vs 68 years (IQR 56-76), p=0.018, Table 1) and were more commonly male (p=0.015), with the average largest brain metastasis also being greater in males (p=0.017, Figure 1a). MBM are known to have a strong propensity towards spontaneous haemorrhage[16], an important cause of morbidity and mortality, and we observed female patients were more likely to experience a clinically significant intracranial haemorrhage (p=0.035, Figure 1b). This remained the case when adjusting for age, *BRAF* status, pre-treatment and performance status (OR 3.6, 95% CI 1.1-11.9, p=0.033). Conversely, SRS and diameter of the largest brain metastasis were not independently associated with haemorrhage. Other relevant demographics, treatments and features of their disease are provided (Table 1).

**Figure 1.**
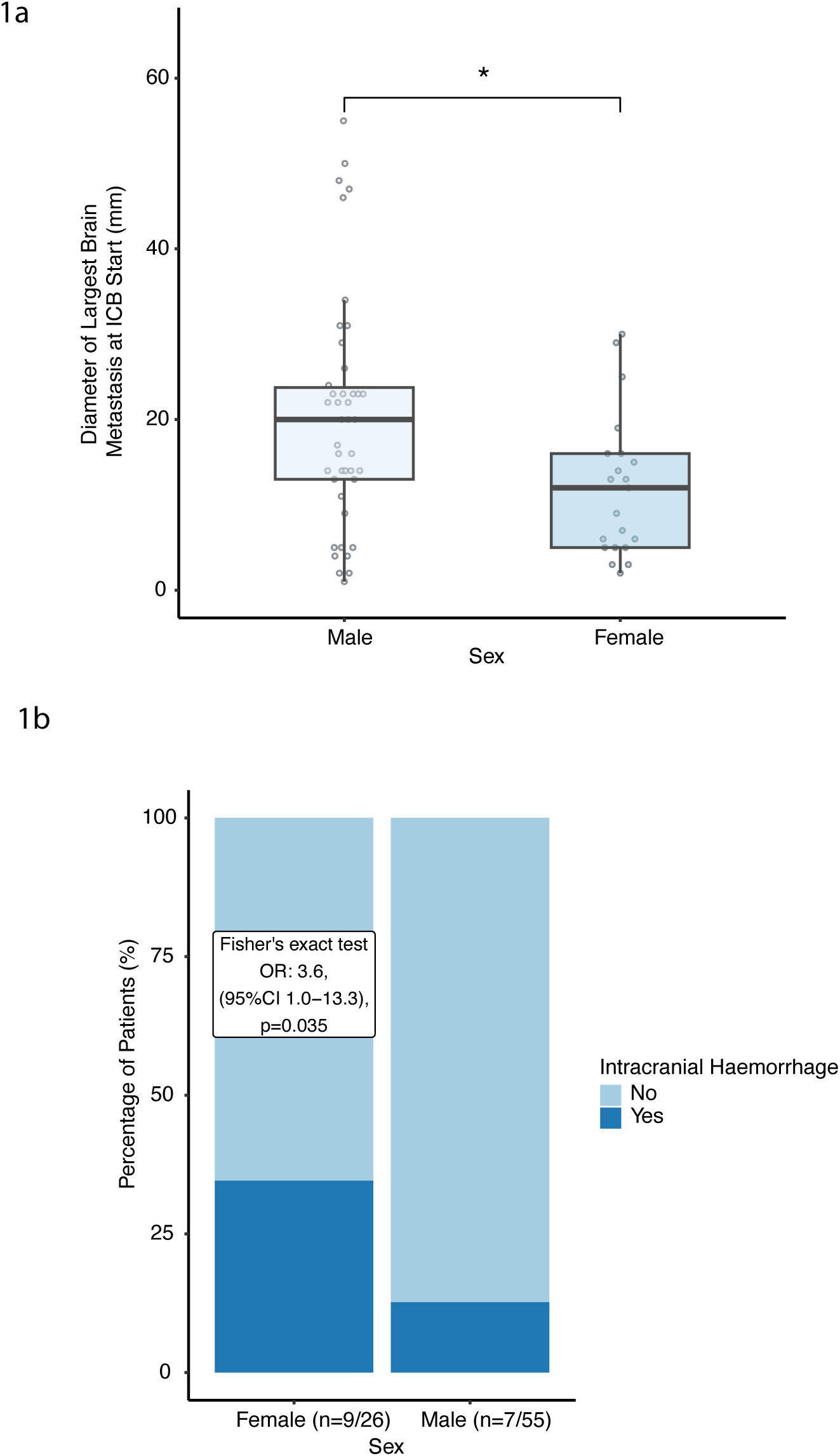
Biological sex associations of MBM. 1a) Diameter of largest brain metastasis at ICB start by sex (p=0.017, Mann-Whitney U test), 1b) Symptomatic intracranial haemorrhage in patients with brain metastases at ICB start, by sex.

**Table 1.**

| Characteristic | Brain Metastases at ICB Start |  | p-value<sup>2</sup> |
| --- | --- | --- | --- |
|  | No<br>N = 226<sup>1</sup> | Yes<br>N = 81<sup>1</sup> |  |
| **Age** | 68 (56, 76) | 62 (53, 72) | 0.018 |
| **Sex** |  |  | 0.015 |
| Male | 118 (52%) | 55 (68%) |  |
| Female | 108 (48%) | 26 (32%) |  |
| **Performance Status** |  |  | 0.058 |
| 0 | 139 (62%) | 40 (49%) |  |
| ≥1 | 87 (38%) | 41 (51%) |  |
| **Treatment** |  |  | <0.001 |
| Ipilimumab and Nivolumab | 132 (58%) | 69 (85%) |  |
| Nivolumab | 20 (8.8%) | 0 (0%) |  |
| Pembrolizumab | 74 (33%) | 12 (15%) |  |
| **BRAF Status** |  |  | <0.001 |
| Wild-type | 158 (70%) | 39 (48%) |  |
| Mutant | 68 (30%) | 42 (52%) |  |
| **Prior treatment with BRAF/MEKi or ICB** |  |  | <0.001 |
| Not pre-treated | 183 (81%) | 50 (62%) |  |
| Pre-treated | 43 (19%) | 31 (38%) |  |
| **MDT Outcome** |  |  |  |
| Not eligible for local treatment | - | 18 (22%) |  |
| Not referred to MDT | - | 20 (25%) |  |
| Recommended local treatment | - | 32 (40%) |  |
| Recommended to proceed with ICB | - | 11 (14%) |  |

<sup>1</sup> Median (Q1, Q3); n (%)<sup>2</sup> Wilcoxon rank sum test; Pearson's Chi-squared test

#### 3.1.1 *BRAF* mutation

Patients with MBM were more likely to have *BRAF*-mutated disease (p*→* 0.001, Table 1). Moreover, patients with *BRAF*-mutated disease and MBM were younger than those without. Median age was 69 years in the *BRAF* wild-type cohort and 60 years in the *BRAF* mutant cohort (p*→* 0.001). This is in keeping with the *BRAF* mutation being disproportionately found in younger patients[14]. Consistent with *BRAF*-mutated disease having a divergent disease trajectory, amongst patients without MBM, excluding those alive and censored before one year of follow-up post ICB commencement (n=206), carriage of a *BRAF* mutation was additionally associated with increased likelihood of developing *de novo* brain metastases whilst receiving ICB treatment (Table 3), an observation robust to multivariable analysis with binominal logistic regression (OR 2.5, 95% CI 1.1-5.9, p=0.036). In line with evidence from retrospective analyses and trials such as DREAMseq[15], practice has shifted from starting BRAF/MEKi as first line therapy in patients with MBM. Nonetheless, within the NHS, combination ipilimumab-nivolumab and relatlimab-nivolumab are only licensed for patients with asymptomatic MBM [17, 18]. Thus a subset of patients with *BRAF*-mutated disease still commence BRAF/MEKi as first line therapy. Reflecting this, prior treatment with BRAF/MEKi or ICB is overrepresented in the MBM cohort (Table 1). Amongst the BRAF/MEKi pretreated patients, 14/39 had brain metastases upon initiation of BRAF/MEKi treatment, with 22/39 having brain metastases when commencing ICB, reflecting the brain forming a frequent site of *de novo* metastasis in this patient group. Amongst these patients the median progression-free survival prior to switching to ICB was 6 months.

### 3.2 Determinants of survival

For patients with a known MBM at the time of commencing ICB treatment, in spite of males having on average larger metastases, we found no association between either age or sex and OS (Table 2). A weak sex association was noted for the development of brain metastases after ICB treatment started however, with females nominally more likely to develop *de novo* brain metastases (Table 3). Applying multivariable analysis, OS was similar across patients with and without MBM from the time they commenced ICB treatment (Figure 2a). This is in contrast to patients with MM in OxCITE who had hepatic metastases, a frequent occurrence described to be associated with poor prognosis [19, 20]. As per other studies, we observed that patients with one or more hepatic metastases have a markedly shorter OS, with median OS of 13 months, compared to 24 months for those without, a finding robust to multivariable analysis (HR 1.7, 95% CI 1.2-2.4, p=0.0019, Figure 2b).

**Figure 2.**
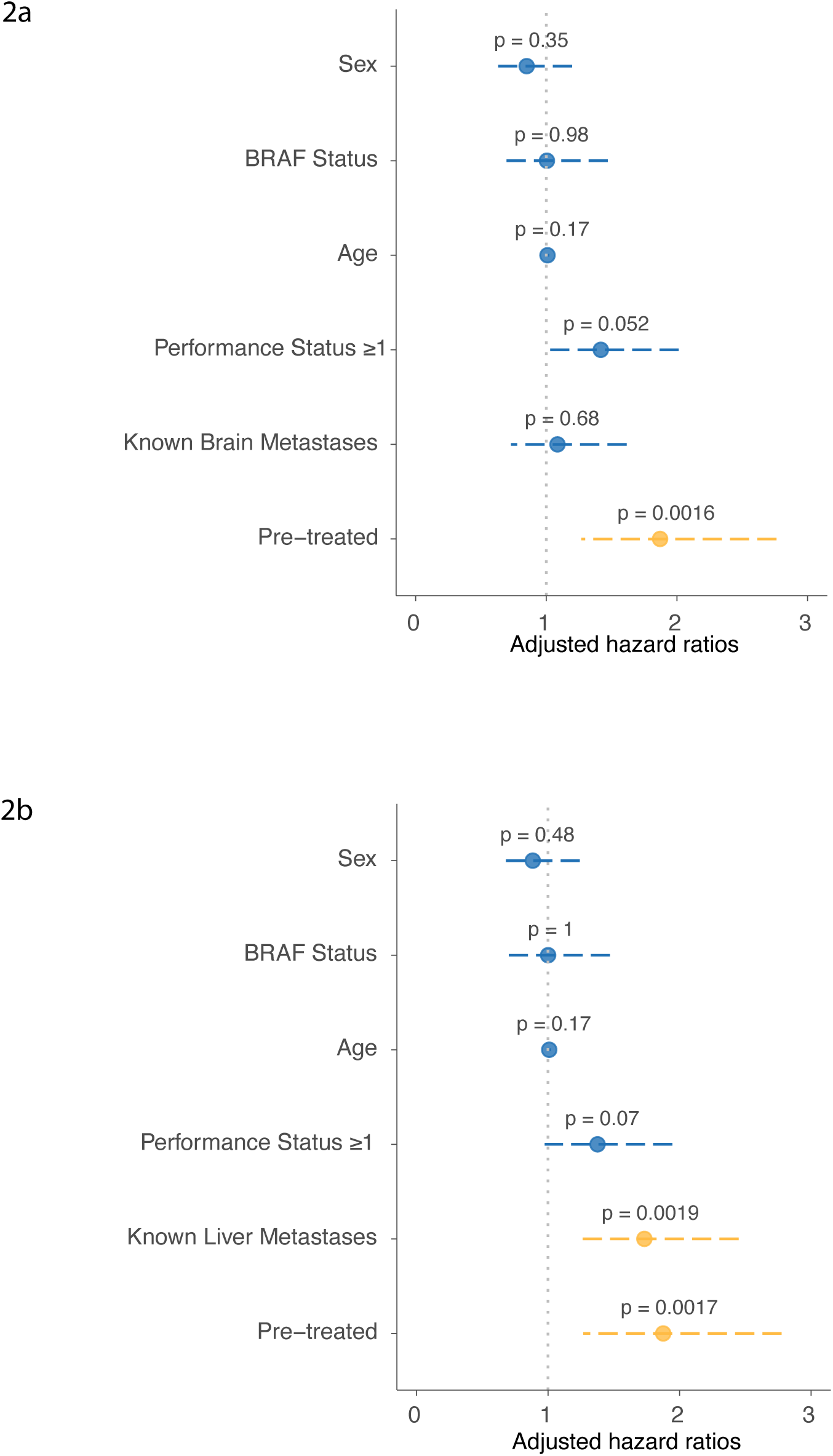
Multivariable cox analysis for OS, patients with metastatic melanoma. 2a) Including brain metastases at ICB start, 2b) Including hepatic metastases at ICB start

**Table 2.**

| Characteristic | Univariable |  |  |  |  | Multivariable |  |  |
| --- | --- | --- | --- | --- | --- | --- | --- | --- |
|  | N | Event N | HR | 95% CI | p-value | HR | 95% CI | p-value |
| Age | 81 | 36 | 1.00 | 0.97, 1.03 | 0.95 |  |  |  |
| Sex |  |  |  |  |  |  |  |  |
| Male | 55 | 27 | — | — |  |  |  |  |
| Female | 26 | 9 | 0.57 | 0.27, 1.21 | 0.14 |  |  |  |
| BRAF Status |  |  |  |  |  |  |  |  |
| Wild-type | 39 | 13 | — | — |  | — | — |  |
| Mutant | 42 | 23 | 2.12 | 1.07, 4.21 | **0.031** | 1.61 | 0.76, 3.43 | 0.21 |
| Prior treatment with BRAF/MEKi or ICB |  |  |  |  |  |  |  |  |
| Not pre-treated | 50 | 19 | — | — |  | — | — |  |
| Pre-treated | 31 | 17 | 1.94 | 1.00, 3.74 | **0.048** | 1.74 | 0.84, 3.60 | 0.13 |
| Performance Status ≥1 |  |  |  |  |  |  |  |  |
| 0 | 40 | 20 | — | — |  |  |  |  |
| ≥1 | 41 | 16 | 0.92 | 0.47, 1.77 | 0.79 |  |  |  |
| ICB Treatment |  |  |  |  |  |  |  |  |
| Combined | 69 | 27 | — | — |  |  |  |  |
| Single Agent | 12 | 9 | 1.85 | 0.86, 3.94 | 0.11 |  |  |  |
| Local Treatment |  |  |  |  |  |  |  |  |
| No Local Treatment | 45 | 23 | — | — |  |  |  |  |
| Other Palliative RT | 5 | 3 | 1.09 | 0.33, 3.63 | 0.89 |  |  |  |
| Resection Only | 3 | 1 | 0.44 | 0.06, 3.27 | 0.42 |  |  |  |
| SRS and Resection | 9 | 3 | 0.51 | 0.15, 1.69 | 0.27 |  |  |  |
| SRS Only | 19 | 6 | 0.55 | 0.22, 1.36 | 0.19 |  |  |  |
| Symptoms at First Presentation |  |  |  |  |  |  |  |  |
| Absent | 40 | 16 | — | — |  |  |  |  |
| Present | 41 | 20 | 1.47 | 0.76, 2.84 | 0.25 |  |  |  |
| On Steroids for Brain Metastases at ICB Start |  |  |  |  |  |  |  |  |
| Not Prescribed | 60 | 21 | — | — |  | — | — |  |
| Prescribed | 21 | 15 | 2.54 | 1.30, 4.96 | **0.006** | 2.56 | 1.30, 5.08 | **0.007** |
| Antiepileptic Medications |  |  |  |  |  |  |  |  |
| Not Prescribed | 55 | 21 | — | — |  |  |  |  |
| Prescribed | 26 | 15 | 1.74 | 0.89, 3.37 | 0.10 |  |  |  |
| Diameter of Largest Brain Metastasis at ICB Start | 63 | 28 | 1.02 | 0.99, 1.05 | 0.18 |  |  |  |
| Symptomatic Intracranial Haemorrhage |  |  |  |  |  |  |  |  |
| Absent | 65 | 27 | — | — |  |  |  |  |
| Present | 16 | 9 | 1.71 | 0.80, 3.66 | 0.17 |  |  |  |
| Abbreviations: CI = Confidence Interval, HR = Hazard Ratio |  |  |  |  |  |  |  |  |

Abbreviations: CI = Confidence Interval, HR = Hazard Ratio

**Table 3.**

| Table 3 |  |  |  |
| --- | --- | --- | --- |
| Characteristic | Developed Brain Metastases within 1 Year of Commencing ICB |  | p-value<sup>2</sup> |
|  | No<br>N = 175<sup>1</sup> | Yes<br>N = 31<sup>1</sup> |  |
| Age | 68 (58, 76) | 59 (46, 74) | 0.067 |
| Sex |  |  | 0.028 |
| Male | 94 (54%) | 10 (32%) |  |
| Female | 81 (46%) | 21 (68%) |  |
| Treatment |  |  | 0.8 |
| Combined | 103 (59%) | 19 (61%) |  |
| Single Agent | 72 (41%) | 12 (39%) |  |
| BRAF Status |  |  | 0.010 |
| Wild-type | 130 (74%) | 16 (52%) |  |
| Mutant | 45 (26%) | 15 (48%) |  |
| Prior treatment with BRAF/MEKi or ICB |  |  | 0.6 |
| Not pre-treated | 140 (80%) | 26 (84%) |  |
| Pre-treated | 35 (20%) | 5 (16%) |  |
| <sup>1</sup> Median (Q1, Q3); n (%) |  |  |  |
| <sup>2</sup> Wilcoxon rank sum test; Pearson's Chi-squared test |  |  |  |

When patients were secondarily analysed according to presence of a *BRAF* mutation, patients with *BRAF* wild-type disease MBM were again not found to be negatively prognostic (Figure 3a). In contrast, across all patients with *BRAF*-mutated disease the presence of MBM was associated with significantly reduced survival compared to those without (Figure 3b). We considered whether this may be due to BRAF/MEKi pretreatment, as pretreated patients had significantly worse overall survival than *BRAF*-mutant patients who did not receive upfront BRAF/MEKi therapy (HR 2.0, 95% CI 1.2-3.6, p=0.014). The reduction in survival with MBM in the *BRAF*-mutant group remained significant when adjusted for pre-treatment in multivariable analysis (HR 2.0, 95% CI 1.1-3.6, p=0.024) but was not observed when pre-treated patients were removed from the cohort altogether (Figure 3c), although this reduces power. Conversely, a *BRAF*-associated difference was not observed in the group with hepatic metastases and there was no difference in survival between *BRAF* mutant and wild-type patients across the entire metastatic cohort (n=307, HR=1.1, 95% CI 0.75-1.5, p=0.72) or in the brain metastasis group in multivariable analysis (Table 2).

**Figure 3.**
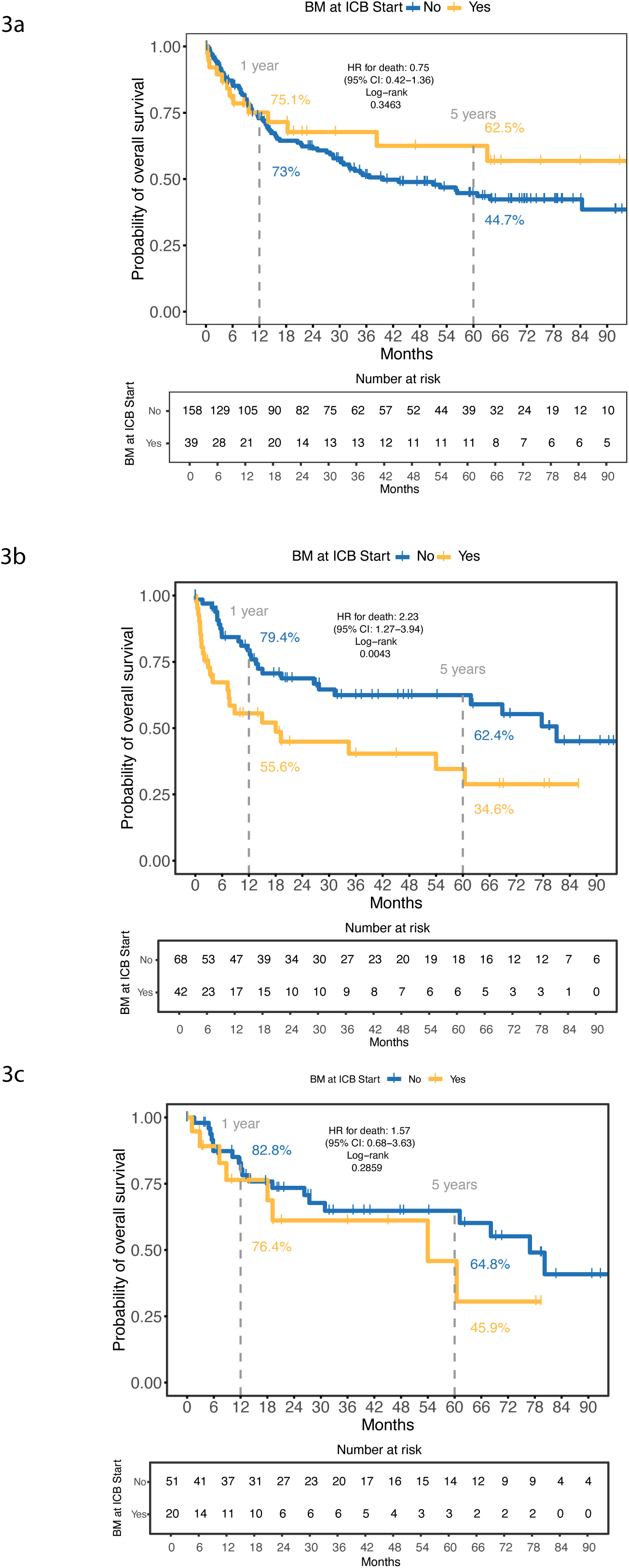
**Association between presence of brain metastases at ICB start with OS, *BRAF* wild-type vs *BRAF* mutant patients**: 3a) Kaplan-Meier curve for OS according to the presence of brain metastases at ICB start, *BRAF* wild-type patients, 3b) Kaplan-Meier curve for OS according to the presence of brain metastases at ICB start,*BRAF* mutant patients, 3c) Kaplan-Meier curve for OS according to the presence of brain metastases at ICB start,*BRAF* mutant patients who had not been pre-treated

Twelve patients who had brain metastases on commencing ICB treatment did not meet the eligibility criteria for combination therapy with ipilimumab and nivolumab. Therefore, they received single-agent treatment with pembrolizumab. Interestingly, their survival was not found to be significantly worse (Table 2), although we note the small number of patients here limits power. There was however no significant difference in the size or number of brain metastases between the single agent and combination ICB groups. On further analysis, only one patient who received single agent ICB and lived beyond 12 months from treatment initiation had not received radiotherapeutic or surgical intervention.

### 3.3 Additional Treatments

Patients who were recommended local treatments by the neuro-oncology multidisciplinary team (MDT) (Table 1) did not reach median survival, with a 5-year survival rate of 55%. Those who were deemed not to require local treatment had similar survival with a median survival of 63 months. This is in contrast to those who were deemed ineligible for local treatments (median OS 4.8 months) or not referred due to heavy disease burden (median OS 15 months). There was a nominal association between SRS and increased toxicity requiring steroid treatment in binomial logistic regression, however this did not carry through to a time-dependent model due to shorter OS in the group who did not receive local treatment.

### 3.4 Other factors

Exploring other potentially negatively prognostic factors amongst patients, 41 patients were symptomatic at their first presentation of MBM, but these had no worse OS (Table 2). However, 22 of these patients had local treatment with SRS and/or resection prior to commencing ICB which may explain this. We identified 26 patients receiving anti-seizure treatment, a proxy of symptoms, but found no worsening of survival in these patients (Table 2). Only four patients were identified with leptomeningeal metastases which was associated with very short OS (median 0.48 months). Finally, 21 patients in this cohort commenced ICB treatment while receiving corticosteroids. This group consisted of 12 patients who had not completed their steroid weaning following a symptomatic presentation and 9 who had not completed their steroid weaning following SRS. Overall, these patients had worse survival than those who did not receive steroids (Figure 4a, Table 2). Whilst 8/21 patients receiving steroids at ICB initiation survived beyond 12 months, we noted 5 of these patients had received steroids as part of their SRS treatment regimen. Although controlling for receipt of SRS, in conjunction with age, sex and *BRAF* status did not remove the negative association of steroids (HR for death 3.1, 95% CI 1.5-6.2, p=0.0014), subgroup analysis of patients in receipt of steroids outside of the context of SRS indicated dire outcomes, with median survival of 3.6 months (Figure 4b). Conversely, we observed that the 26 patients who had received steroids but stopped *prior* to commencing ICB had no evidence of worse survival compared to patients who did not receive steroids (adjusted HR 0.49, 95% CI 0.18-1.34, p=0.17) and patients without brain metastases (HR 0.57, 95% CI 0.26-1.24, p=0.15). However, it should be noted that 19 of these patients received SRS and 2 received other forms of palliative radiotherapy.

**Figure 4.**
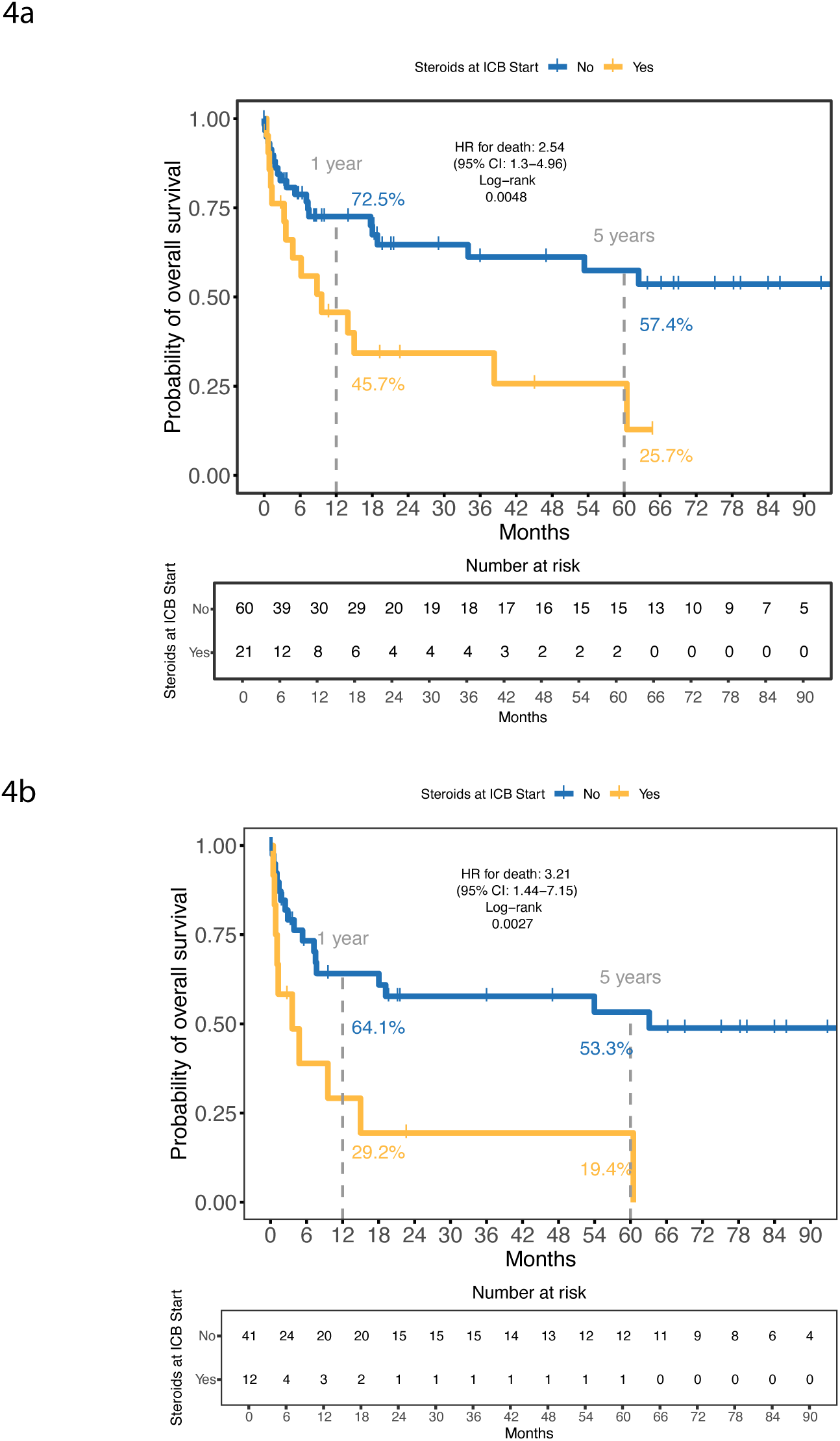
Association between steroid treatment at ICB initiation and outcomes in patients with melanoma brain metastases. 4a) Kaplan-Meier curves for OS according to steroid treatment status at ICB start across all patients with MBM, 4b) Kaplan-Meier curves for OS according to steroid treatment status at ICB start in patients with MBM who did not receive SRS

### 3.5 Peripheral markers of brain metastases

There are currently few peripheral biomarkers of CNS disease. Therefore, some possibilities were explored in our cohort. Increased neutrophil-lymphocyte ratio (NLR) is a recognised adversely prognostic indicator in metastatic melanoma[21]. On multivariable linear regression including age, sex, *BRAF* status and pre-treatment, patients with brain metastases had a 1.3-fold increase in NLR compared to those without (n = 79/299, 95%CI 1.1-1.5, p=0.0038). Similarly, patients with liver metastases were found to have a 1.2-fold increase in NLR compared to those without (n= 80/299, 95% CI 1.1-1.4, p=0.0093), indicating that the relationship was not specific to brain metastases. Both increasing NLR and liver metastatic disease independently associated with worse OS in a multivariable model, while brain metastases did not (Figure 5a). Conversely, baseline LDH was not elevated among patients with MBM (n = 54/190, 1.14-fold change, 95% CI 0.96-1.36, p=0.13), whereas it was markedly elevated in those with liver metastases (n = 57/190, 1.41-fold change, 95%CI 1.20-1.64, p*→* 0.001). These data suggest that elevated LDH is more specifically associated with liver metastases than elevated NLR, which may be a broader marker of disease burden.

**Figure 5.**
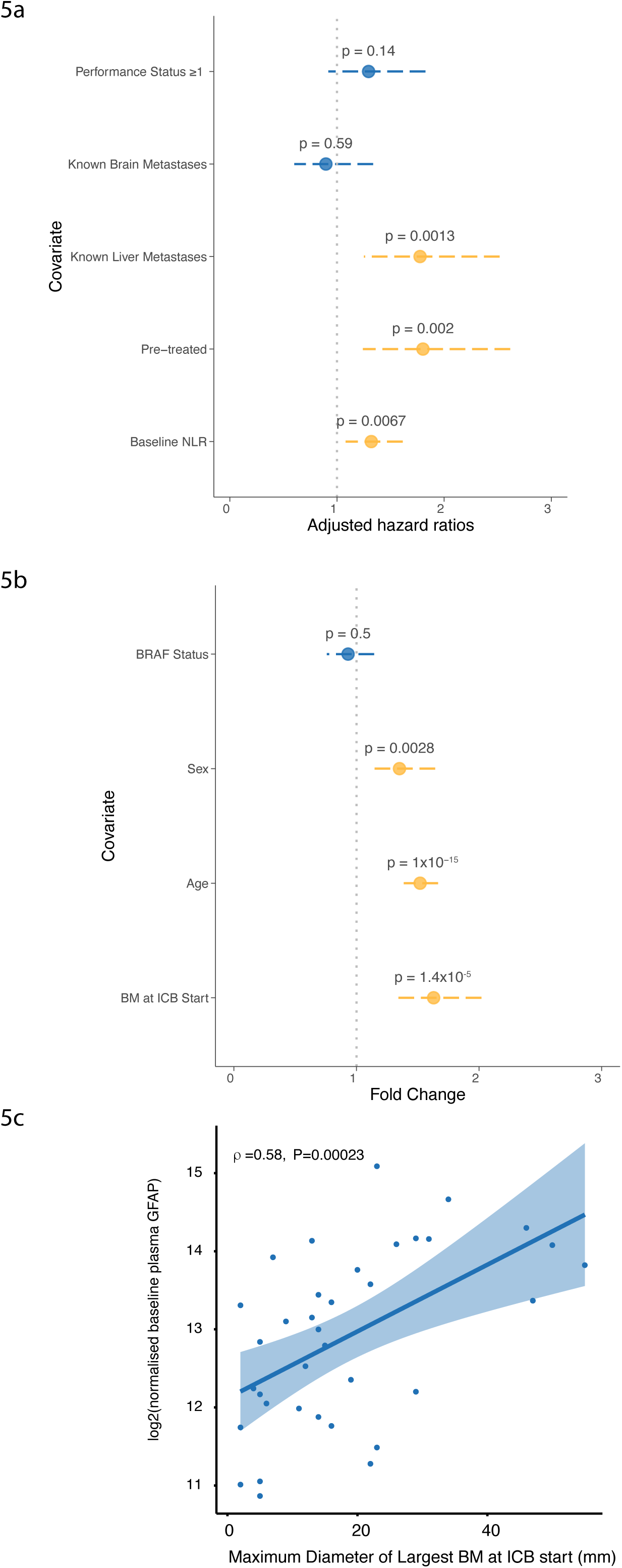
**Association between peripheral biomarkers, survival and intracranial disease burden**, 5a) Multivariable cox analysis for OS, patients with metastatic melanoma, 5b) Multivariable linear regression for GFAP level at ICB start, patients with metastatic melanoma, 5c) Association between baseline plasma GFAP level and diameter of the largest brain metastasis at the start of ICB treatment

Finally, we sought to explore whether markers known to be elevated in the context of neurological disease may show differences in the context of MBM. In patients where baseline pre-treatment plasma samples were available (n=152 patients), we assessed glial fibrillary acidic protein (GFAP) levels, a protein that is markedly elevated in traumatic brain injury and Alzheimer’s disease [22, 23], using a nucleic acid-linked immuno-sandwich assay. Intriguingly, after adjusting for age, sex and *BRAF* status, we found plasma GFAP to be increased in patients with brain metastases at baseline (Figure 5b). Moreover, GFAP levels were positively correlated with maximum diameter of the largest brain metastasis at ICB start (n=36, Spearman’s rho=0.58, P=2.3×10*^→^*^4^, Figure 5c), an observation that remains robust to correction for age and sex (P=1.5×10*^→^*^4^).

## 4 Discussion

In this study of prospectively recruited patients with melanoma we found that the presence of brain metastases at the initiation of ICB treatment was not associated with poorer OS. Our experience is in keeping with observations from the two main studies exploring the role of ICB in MBM, Checkmate 204 and ABC. These reported intracranial response rates in asymptomatic patients of 53.5% and 51% respectively[10][11]. Correspondingly, 36 month OS in the asymptomatic arm of Checkmate 204 was 71.9%, whilst 7 year OS in ABC ipilimumab/nivolumab recipients was 48%. Importantly, our dataset has a similar number of patients to these studies and reflects real-world experiences in the NHS, where the criteria for entering the good prognostic arms of Checkmate 204, which precluded steroid use, and ABC, which excluded patients taking steroids *→*14 days before ICB, were not always met. A key difference between our study and ABC was the lack of observed survival difference between the single agent and combined cohorts (48% vs 26% in ABC). This could be explained by characteristics of our single agent cohort, where 6/12 had OS *>*12 months. Of these 6 longer-term survivors, 5 had received local treatments for their brain metastases, and thus had other contributory causes for local disease control and improved survival outcomes. Broadly, we did not find that symptomatic initial presentation in isolation was sufficient to define poorer prognosis.

In our cohort we find that men were more likely to present with MBM, although women were more likely to have an associated haemorrhage over their treatment course. Irrespective of these findings, we found no association between biological sex and OS in patients with MBM. Similarly, we found that *BRAF* mutated disease was significantly more likely to present with brain metastases. Amongst those with MBM, patients whose tumour carried a *BRAF* mutation had marginally poorer OS than those with wild-type *BRAF* status, although this finding resolved in multivariable analysis. When restricting comparisons to patients with WT *BRAF*, survival was not associated with the presence of MBM. Conversely, amongst patients with *BRAF* mutated melanoma, those with MBM had inferior survival. A significant subset of these patients (22 of the 42 with *BRAF*-mutant MBM) had progressive disease on BRAF/MEKi prior to ICB and it is therefore likely some of this signal corresponds to a pre-treatment effect, with pre-treatment more likely in those with MBM. Further studies across other datasets will be useful to clarify these observations.

Our analysis revealed that continued steroid usage at ICB initiation in patients with MBM - given for persistent symptomatic disease or as a short-weaning course in the context of SRS treatment - was strongly negatively prognostic. These patients demonstrated markedly reduced OS, an effect that did not resolve upon controlling for receipt of SRS. Nonetheless, sub-group analysis of patients receiving steroids outside of the context of SRS at the initiation of ICB highlighted particularly poor outcomes, with median survival of 3.6 months. These findings align with reduced objective response of MBM in steroid recipients within the symptomatic arm of Checkmate 204 [10], and poorer OS amongst steroid recipients of combination anti-PD-1/anti-CTLA-4 ICB in registry studies[24]. Whilst the European Society for Medical Oncology guidelines [25, 26] recommend consideration of ICB treatment in patients with symptomatic CNS disease, our findings suggest that outside of the context of locally directed treatment, benefits are minimal if steroids cannot be discontinued prior to treatment. These findings are consistent with steroid-induced immunosuppression leading to reduced ICB efficacy and would support clinicians weaning steroids prior to ICB initiation where possible.

Whereas early studies of SRS for MBM demonstrated median OS around 6-9 months[27–29], our data show that in conjunction with ICB, patients requiring SRS can derive long-term benefits similar to those with disease not-requiring local intervention. We also noted that SRS may potentially contribute to the development of irAEs, with increased rates of these in SRS recipients. However, this is a preliminary observation and we were not able to formally confirm this using competing risk models due to potential survivor bias in those receiving SRS, increasing the period of time over which an irAE may develop. Nonetheless, as the evidence now strongly favours upfront ICB in asymptomatic patients, the potential relationship between SRS and irAEs warrants further examination.

Given the propensity of melanoma to brain metastases and the relatively good responses of these to both ICB and SRS, brain imaging forms a core component of surveillance follow-up for patients with Stage IIB disease and above. This places a demand on health services and lags between CNS disease and initiation of treatment are inevitable, especially where imaging is biannually. This translates into a requirement to develop supportive orthogonal biomarkers of MBM. Here we show, as far as we are aware, the first demonstration of a significant association between plasma GFAP and the presence of brain metastases in melanoma. Not only are plasma GFAP levels elevated in patients with MBM, but they also correlate with the magnitude of CNS disease burden. Whilst GFAP is a well recognised marker of primary brain tumours, there is limited literature regarding its utility in the context of metastastic disease. A recent retrospective analysis of a historical cohort of patients with breast cancer showed both significant elevations in those with brain metastases, as well as prognostic utility[30]. In our data, although there were associations with disease burden, we did not note an on-treatment predictive value in this small cohort. Nonetheless, our findings suggest that, taking into account known age and sex associations with GFAP[31], serial analysis of GFAP may have utility as part of routine surveillance for high risk melanoma by providing an early adjunctive indicator of MBM in the context of deviation from personal normal ranges. Further prospective studies should be performed to demonstrate a potential role for this in standard practice.

This study has various limitations. First, although survival and toxicity data were collected prospectively, in-depth analysis of brain metastases was conducted retrospectively. This has resulted in some missing data, such as the total volume of brain metastases and whether symptoms were present at the start of ICB, as these were not available in routine clinical data. Likewise, it was not possible to consistently differentiate between prophylactic and symptomatic use of anti-seizure drugs, which may explain the lack of survival difference. The exact dose of corticosteroids patients were receiving was also not always available, precluding exploration of dosage effects. Because imaging for patients with brain metastases varies in timing according to symptoms and other interventions, objective response data were not consistently assessed, thus we focus on survival as the primary endpoint. Finally, the OxCITE cohort is selected for patients who are receiving ICB treatment and will exclude those who are deemed ineligible for ICB due to fitness or other precluding co-morbidities.

Prior to the introduction of ICB, the proportion of melanoma deaths attributable to intracranial disease was estimated at 20–54%[4, 32, 33]. The advent of standard-of-care ICB has changed the paradigm of melanoma brain metastases as much as it has changed that of advanced melanoma in general. Patients are able to mount deep and sustained responses even with significant CNS disease, which can also be supported by local control. Indeed, hepatic metastases are associated with poorer response rates and outcomes than MBM[19, 20]. Our data build on those of others to suggest that in the context of treatment with ICB, the M1d classification of brain metastases no longer carries the prognostic weight it once did.

## Data Availability

Anonymised relevant data and scripts used in this study are available upon reasonable request to the authors.

